# Comparison of Efficacy, Safety, and Survival Outcomes of Anticoagulation and Antiplatelet Strategies Post Liver Transplantation: A Systematic Review and Network Meta-Analysis

**DOI:** 10.1101/2025.10.07.25337027

**Authors:** Christen En Ya Ong, Rachel Si Yin Wong, Yihan Zhang, Yi Da Chua, Yu Ze Ng, Jia Hao Law, Alfred Kow

## Abstract

**Background:** Liver transplantation (LT) is the definitive treatment for end-stage liver disease, but remains complicated by thrombotic events such as hepatic artery and portal vein thrombosis. This network meta-analysis compares the efficacy and safety of different antithrombotic strategies post-LT, as well as dosing and duration to inform individualized, evidence-based management.

**Methods:** Medline and Embase were searched to 22 November 2024 for RCTs and cohort studies reporting outcomes of postoperative antithrombotic methods in liver transplant patients. Random-effects frequentist network meta-analysis pooled odds ratios (OR), 95% confidence interval (CI) values, and P-score to rank the antithrombotic medications.

**Results:** Among 18 studies involving 8,856 patients, aspirin (OR: 0.30, 95%CI: 0.18 – 0.51, p=0.04) and UFH (OR: 0.31, 95%CI: 0.12 – 0.84, p<0.01) use were associated with lowest overall thrombotic risk relative to the control group. Aspirin was ranked the highest for HAT prevention (P-score = 0.88) and had no significant bleeding risk. Low molecular weight heparin (LMWH) was ranked highest for DVT prevention (P-score = 0.67) but also second for worst associated bleeding risk (P-score = 0.43) after VKA (OR: 3.53, 95%CI: 1.86 – 6.71, p<0.01, P-score = 0.01). LMWH also ranked worse than control in prevention of PVT. DOACs were ranked first for the lowest associated bleeding risk and third for reduction in overall thrombotic risk. None of the antithrombotic medications showed any significant association with overall patient mortality.

**Conclusions:** Aspirin remains a mainstay of arterial thromboprophylaxis. DOACs appear promising for thromboprophylaxis post-LT, with the lowest bleeding risk. Targeted rather than routine prophylaxis, guided by individual risk profiles, likely maximises post-LT outcomes.

## INTRODUCTION

Liver transplantation (LT) remains the definitive treatment modality for patients with end stage liver disease, a deadly condition which accounts for 1.3 million deaths worldwide annually^1,2^. A major complication post-LT is the occurrence of hepatic artery thrombosis (HAT) or portal vein thrombosis (PVT), which can result in complications like graft failure and giving rise to significant morbidity post-operation^3,4^. Arterial and venous thromboses post-LT are clinically distinct conditions, both associated with poor patient outcomes/complications but different management strategies^3^. However, paradoxically, despite the necessity to manage these thrombotic events, the immediate post-operative period is marked by significant risks of bleeding events due to pre-existing coagulopathy^5^. This is also contributed by the delayed normalization of the coagulation profile as the transplanted liver gradually resumes its synthetic functions post operation^6,7^.

Despite the aforementioned risks, there remains no well-established consensus for the use of various types of antithrombotic medications in the post operative thromboprophylaxis of liver transplant recipients worldwide. While numerous studies compare the efficacy and safety of various anticoagulation or antiplatelet measures, the findings across studies for certain medications are inconsistent and a comprehensive review that quantitatively summarises and compares the medications in terms of efficacy in preventing different thrombotic outcomes and safety in terms of risk of complications has yet to exist. Different thromboprophylactic medications have been noted to be more efficacious at preventing different types of thromboses, with antiplatelets such as aspirin recommended for arterial thromboses and anticoagulants including heparins for venous thromboses, but the exact choice of drug, duration and dosage varies widely between practitioners. In addition, no meta-analysis presently includes a comparison between DOACs and conventional agents such as heparin and aspirin, likely due to the novelty of these agents^8^. The insufficient standardisations have led to significant disparities in clinical practice, with treatment modalities relying upon local protocols, clinical judgements and anecdotal experience. Such variability highlights the need for evidence-based strategies to optimize the delicate balance between preventing thrombotic complications and minimizing bleeding risk in this specific patient population.

This paper aims to explore the challenges and differences surrounding anticoagulant and antiplatelet use in liver transplant recipients, with a focus on evaluating the current evidence, identifying gaps in knowledge, and proposing directions for future research to guide clinical decision-making for the management of liver transplant patients. The findings of this network meta-analysis will help advise transplant surgeons on their selection of thromboprophylaxis type tailored to the individual patient’s needs, helping to reduce complications and maximise patient outcomes, and also inform the direction of future studies on post-LT thromboprophylaxis.

## METHODS

This network meta-analysis was performed in accordance with the Preferred Reporting Items for Systematic Reviews and Meta-Analysis (PRISMA) statement^9^. The study protocol is available online and is registered with PROSPERO (CRD420251113462).

### Search strategy

The Medline and Embase electronic databases were searched from inception to 1 May 2025 for articles relating to the use of any antithrombotic method in LT patients. Predefined search terms included but were not limited to “liver transplantation”, “anticoagula$”, “antiplatelet$”, and the complete search strategy can be found in Supplemental Figure 1. The reference lists of retrieved studies were also manually sieved for additional relevant studies not indexed in the databases, to ensure a comprehensive search.

### Study selection and extraction

Four authors (RSYW, YZ, YDC, YZN) in blinded pairs systematically screened all articles to identify articles for full-text review, which were further screened for inclusion in data extraction. Removal of article duplicates was handled by Endnote X9. For the network meta-analysis, we included articles that compared post-operative outcomes between at least two of the following: aspirin, direct oral anticoagulants (DOACs), unfractionated heparin (UFH) only, vitamin K antagonists (VKA) such as warfarin (with or without heparin bridging in the immediate postoperative period), low molecular weight heparin (LMWH), prostaglandins and analogues, or no antithrombotic medications used in post-LT patients. We included studies that described the patient, donor, and operative characteristics and outcomes in adult patients undergoing LT with the postoperative use of anticoagulant or antiplatelet medications. Pediatric, animal and non-English studies were excluded, as well as case reports and conference abstracts due to their typically incomplete reporting, high risk of bias, and limited impact on review conclusions, as supported by previous research^10,11^.

Data was extracted by 2 blinded pairs of authors (RSYW, YZ, YDC, YZN) for internal reliability and consistency, and a fifth author (CEYO) was involved to resolve discrepancies. Study characteristics such as year of publication, study duration, mean recipient age, mean MELD score at LT, mean graft weight, mean cold and warm ischemia times were extracted. Intervention characteristics extracted were type of anticoagulant or antiplatelet used, dosage, route of administration and duration of therapy. The primary outcomes of interest for the network meta-analysis were post-LT patient mortality, thrombotic events and bleeding events. Secondary outcomes were split into arterial complications, namely hepatic artery thrombosis (HAT), and venous complications, specifically portal vein thrombosis (PVT) and deep vein thrombosis (DVT). For the purpose of the meta-analysis, mean and standard deviation that were unavailable for continuous outcomes were estimated from the formula provided by Wan et al^12^.

### Statistical analysis and quality assessment

Network meta-analysis was performed using the netmeta package in R 4.4.2. The network meta-analysis was performed using a frequentist approach, allowing for indirect comparison and ranking of PIM methods for each outcome^13^. The meta-analysis was performed in the natural log of odds ratio (lnOR) or hazard ratio (lnHR) and results were exponentiated to obtain the odds ratio (OR) and hazard ratio (HR). Network geometry diagrams were drawn, with the nodes representing thromboprophylaxis methods and the thickness of connecting lines representing the number of studies included. Each treatment comparison was presented with 95% confidence intervals (CI). P-scores were generated to rank the PIM methods for each outcome of interest, where a higher P-score indicated a lower association with said outcome. Network heterogeneity was assessed using I^2^ statistics. Consistency in the network was evaluated using a global design-by-treatment inconsistency model (Q index), and node splitting was performed to identify local inconsistency between direct and indirect comparisons. Publication bias was evaluated for the two primary outcomes (graft failure and patient mortality) by visually inspecting the funnel plot for asymmetry and quantified using the Egger’s test in outcomes with 10 studies or more, as per Cochrane handbook guidelines^14^. Single-arm analysis was performed using the dmetar package in R 4.4.2. All analyses were performed using a DerSimonian-Laird random effects model regardless of heterogeneity scoring, which was quantified using I^2^ and Cochran-Q test.

A subgroup analysis using a simple pairwise approach was planned to compare between different durations of administration for each thromboprophylaxis medication. As only 2 studies for LMWH and UFH each reported duration of use, and regimens used for DOACs and UFH + VKA were too heterogeneous to subgroup meaningfully, this subgroup analysis was only performed for aspirin. Duration of aspirin use was categorized into short-term use (6 months and less) or lifelong. Chi-square test was used as a test for subgroup differences. Differences in dosage and duration of use for the other types of medications were examined qualitatively in the discussion.

To test the robustness of the results, a sensitivity analysis was performed evaluating only RCTs and cohort studies with a low risk of bias. Quality assessment was performed using the Cochrane Risk of Bias 2.0 tool for randomized controlled trials, evaluating randomization process, adherence to intended interventions, missing outcome data, outcome measurement, and selective reporting^15^. Cohort and single-arm studies were evaluated using the Joanna Briggs Institute (JBI) critical appraisal checklist, which assesses study quality based on generalizability of study cohorts, statistical analysis including dealing with confounding variables, reporting of outcomes, and losses to follow-up^16^ A checklist score of 8 and above was determined as a low risk of bias.

## RESULTS

A total of 9,002 articles were retrieved via the literature search in May 2025. After exclusion of 1,048 duplicates, the remaining 7954 articles were screened for eligibility for inclusion. After screening the abstracts and full texts of these articles, 18 articles were included in the analysis^17–34^. Figure 1 shows the study selection process according to PRISMA guidelines. In the network analysis, we used 12 cohort studies and 1 randomized-controlled trial. Supplemental Figure 2 and 3 summarises the characteristics of the studies included, as well as type, dosage and duration of therapy administered and quality of evidence. The overall GRADE score for the evidence is summarised in Supplemental Figure 4.

**Figure 1.**
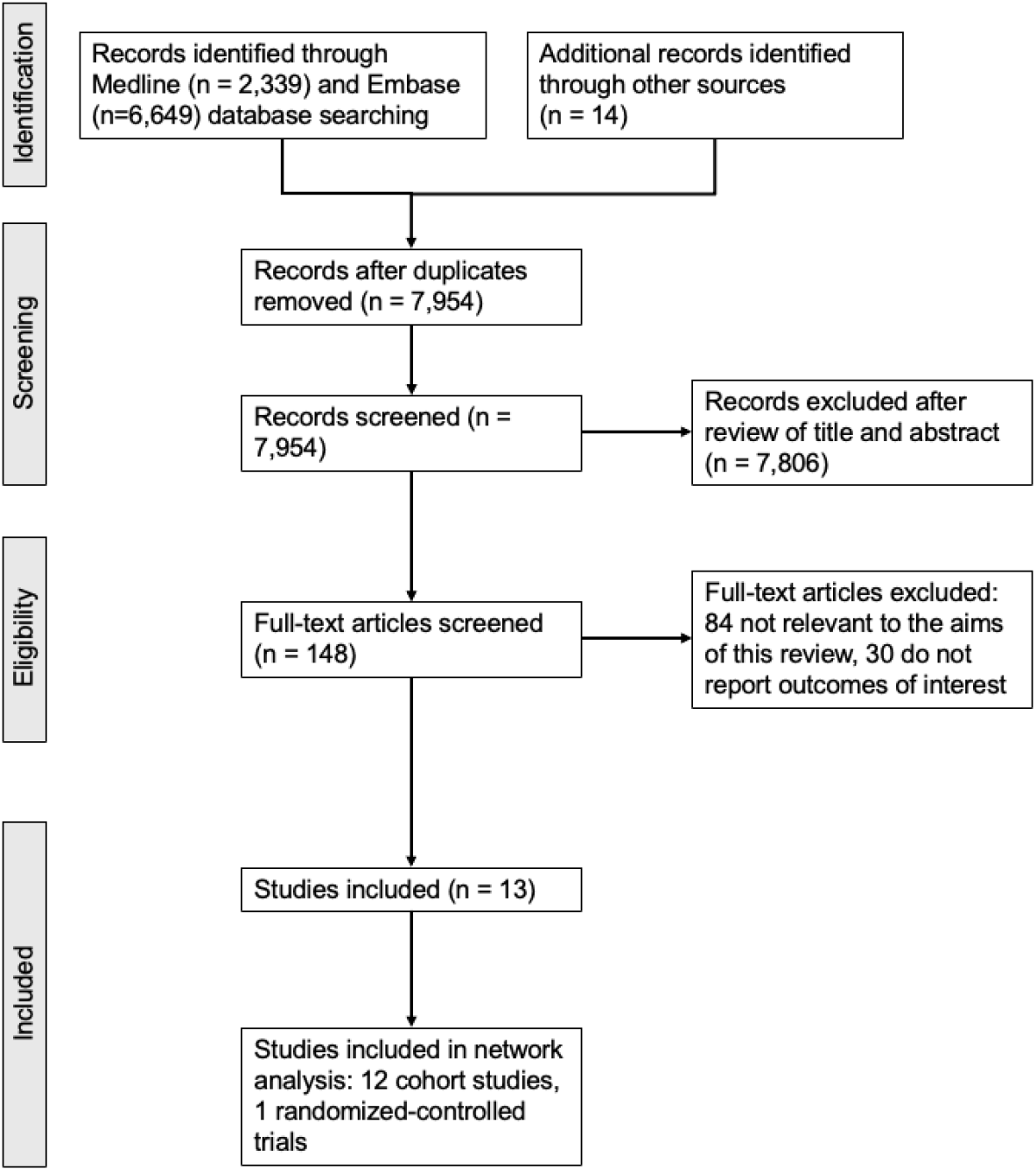
PRISMA flow diagram showing study identification and selection process.

A total of 7,960 patients were included in the meta-analysis. For the network meta-analysis, 5 articles directly compared aspirin and no intervention, 2 compared UFH and no intervention, 4 compared VKA and no intervention, 1 compared LMWH and no intervention,, and 1 compared DOACs and VKA. The baseline characteristics of each patient group is summarized in Table 1.

**Table 1.**
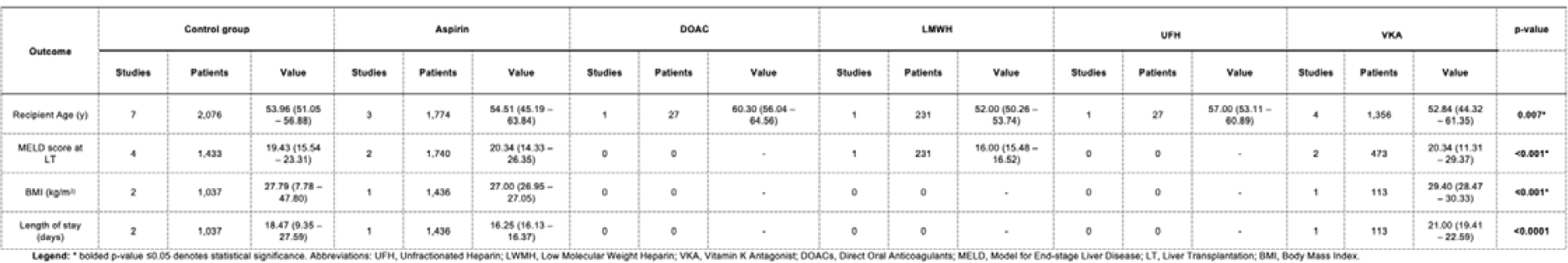
Comparison of the characteristics of the patients of each type of thromboprophylaxis.

### Network meta-analysis

Network meta-analysis ranking the anti-thrombotic agents was performed, and study comparisons for each outcome are represented by the network diagrams in Figure 2. Results of the analysis are summarized in Figure 3 and the ranking of treatments by P-scoring in Table 2.

**Table 2.** P-score ranking from best to worst (1^st^ to 8^th^) for the outcomes of network meta-analysis.

| Outcome | 1st | 2nd | 3rd | 4th | 5th | 6th |
| --- | --- | --- | --- | --- | --- | --- |
| Thrombotic events | Aspirin | UFH | DOAC | VKA | LMWH | Control |
|  | (P = 0.780) | (P = 0.738) | (P = 0.724) | (P = 0.372) | (P = 0.276) | (P = 0.110) |
| Patient mortality | Aspirin | VKA | LMWH | Control | UFH | - |
|  | (P=0.696) | (P=0.635) | (P=0.463) | (P=0.398) | (P = 0.308) | - |
| PVT | Control | LMWH | VKA | - | - | - |
|  | (P = 0.609) | (P=0.498) | (P = 0.393) | - | - | - |
| HAT | Aspirin | VKA | Control | - | - | - |
|  | (P = 0.885) | (P = 0.406) | (P = 0.209) | - | - | - |
| DVT | LMWH | VKA | Control | UFH | - | - |
|  | (P = 0.750) | (P = 0.587) | (P = 0.357) | (P = 0.306) | - | - |
| Bleeding events | DOACs | Control | Aspirin | LMWH | VKA | - |
|  | (P = 0.928) | (P = 0.592) | (P = 0.541) | (P = 0.430) | (P = 0.010) | - |

**Figure 2.**
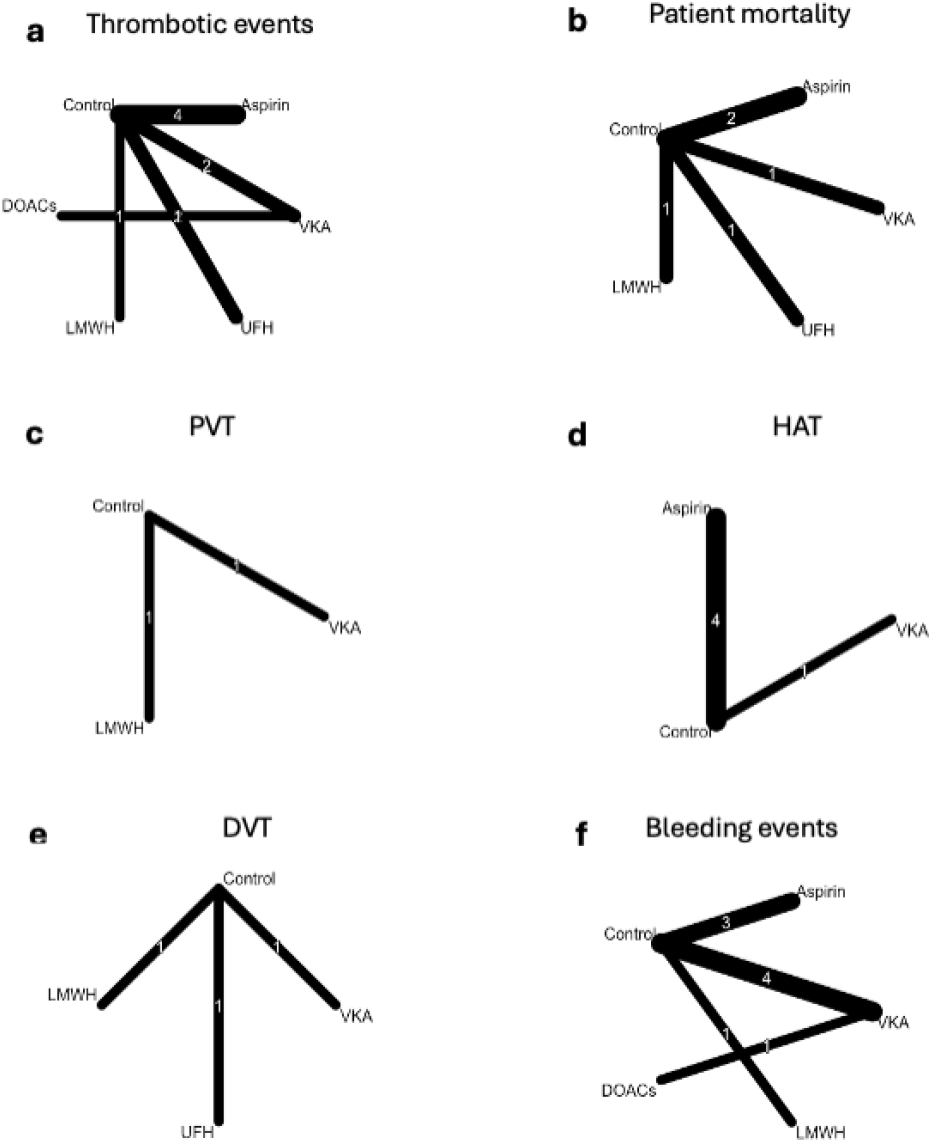
Network diagrams for outcomes of study. Panels are labelled as follows: a) thrombotic events, b) patient mortality, c) portal venous thrombosis (PVT), d) hepatic artery thrombosis (HAT), e) deep vein thrombosis (DVT), f) bleeding events.No chemoprophylaxis group is the reference for all analyses. Abbreviations: OR, Hazard Ratio; CI, Confidence Interval; DOACs, Direct Oral Anticoagulants; UFH, unfractionated heparin; VKA, Vitamin K Antagonist; LMWH, Low Molecular Weight Heparin; PG, Prostaglandins and analogues.

**Figure 3.**
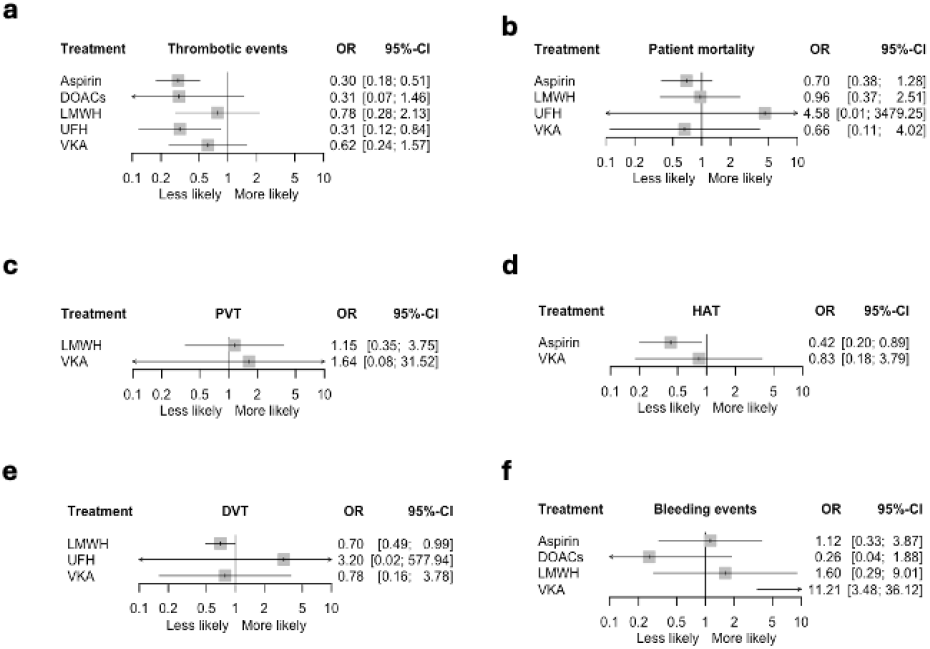
Forest plots for outcomes of study. Panels are labelled as follows: a) thrombotic events, b) patient mortality, c) portal venous thrombosis (PVT), d) hepatic artery thrombosis (HAT), e) deep vein thrombosis (DVT), f) bleeding events. No chemoprophylaxis group is the reference for all analyses. Abbreviations: OR, Hazard Ratio; CI, Confidence Interval; DOACs, Direct Oral Anticoagulants; UFH, unfractionated heparin; VKA, Vitamin K Antagonist; LMWH, Low Molecular Weight Heparin; PG, Prostaglandins and analogues.

#### Thrombotic events

A total of 8 studies involving 7,151 patients were included for the analysis on thrombotic events (Figure 3). Aspirin (OR: 0.30, 95%CI: 0.18 – 0.51, p=0.04) and UFH (OR: 0.31, 95%CI: 0.12 – 0.84, p<0.01) use was associated with a significantly lower risk of postoperative thrombotic events. No significant association was found for DOACs, UFH and LMWH. P-scoring ranked aspirin (P-score = 0.78) first for best outcomes for thrombotic events, followed by UFH (P-score = 0.74), DOACs (P-score = 0.72), VKA (P-score = 0.37), LMWH (P-score = 0.28), and finally control (P-score = 0.11) (Table 2). Moderate heterogeneity (41.2%) was detected according to I^2^. Node splitting did not show evidence of local inconsistency (Supplemental Figure 5). No publication bias was observed (Figure 4) and Egger’s test was non-significant (p=0.51).

**Figure 4.**
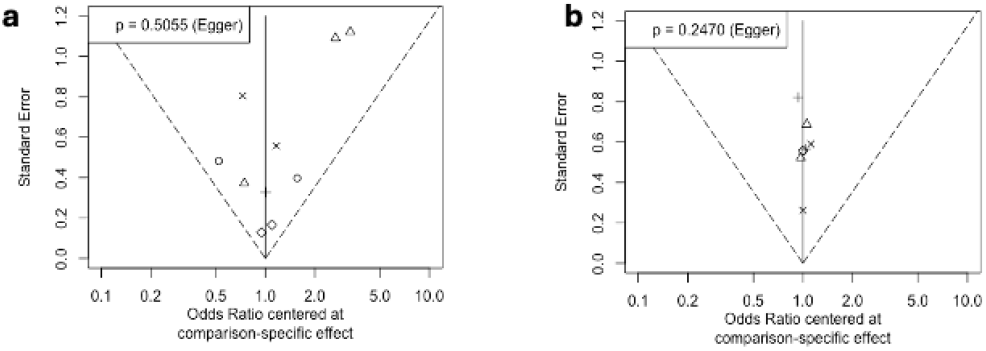
Funnel plots of a) thrombotic events and b) bleeding events. p-values for Egger’s test are displayed on the top left corner.

#### Patient Mortality

A total of 5 studies involving 3,576 patients were included for the analysis on overall patient mortality (Figure 3). No significant associations were found between use of any of the antithrombotic medications studied and patient mortality. P-scoring ranked aspirin (P-score = 0.71) first for best outcomes for patient mortality, followed by VKA (P-score = 0.64), LMWH (P-score = 0.46), control (P-score = 0.40), and finally UFH (P-score = 0.31) (Table 2). No heterogeneity (0%) was detected according to I^2^. Node splitting did not show evidence of local inconsistency (Supplemental Figure 5).

#### PVT

A total of 2 studies involving 697 patients were included for the analysis on overall patient mortality (Figure 3). Both VKA and LMWH showed no association with reduced PVT risk. P-scoring ranked control (P-score = 0.61) first for best outcomes for PVT, followed by LMWH (P-score = 0.50), and VKA (P-score = 0.39) (Table 2).

#### HAT

A total of 5 studies involving 4,412 patients were included for the analysis on overall patient mortality (Figure 3). Aspirin was associated with a lower risk of postoperative HAT (OR: 0.42, 95%CI: 0.20 – 0.89, p<0.01), while VKA use showed no association. P-scoring ranked aspirin (P-score = 0.89) first for best outcomes for HAT, followed by VKA (P-score = 0.41), and control (P-score = 0.21). (Table 2). High heterogeneity (86.8%) was detected according to I^2^.

#### DVT

A total of 3 studies involving 1,693 patients were included for the analysis on overall patient mortality (Figure 3). LMWH (OR: 0.70, 95%CI: 0.49 – 0.99, p = 0.04) was linked to a significantly lower risk of DVT. No significant associations were found between DVT and UFH or VKA use. P-scoring ranked LMWH (P-score = 0.75) first for best outcomes for DVT, followed by VKA (P-score = 0.59), control (P-score = 0.36) and UFH (P-score = 0.31) (Table 2).

#### Bleeding events

A total of 9 studies involving 5,508 patients were included for the analysis on postoperative bleeding events (Figure 3). VKA (OR: 11.21, 95%CI: 3.48 – 36.12, p<0.01) was associated with increased likelihood of postoperative bleeding events. DOACs, LMWH and aspirin showed no significant association. P-scoring ranked DOACs (P-score = 0.93) first for lowest risk of bleeding events, followed by control (P-score = 0.59), aspirin (P-score = 0.54), LMWH (P-score = 0.43), and finally VKA (P-score = 0.01) (Table 2). No heterogeneity (0%) was detected according to I^2^. Node splitting did not show evidence of local inconsistency (Supplemental Figure 5). No publication bias was observed (Figure 4) and Egger’s test was non-significant (p=0.28).

### Subgroup analysis for dosage and duration

Subgroup analysis was performed between high-dose and low-dose aspirin use (Figure 5). Shay et al used high dose aspirin (325mg) daily for a specified 3 month duration. Vivarelli et al, Wolf et al and Oberkofler et al used a daily dose ranging 75-100mg, which we grouped into the low-dose subgroup comprising 3 studies and 3,708 patients. No significant difference in overall thrombotic risk (p=0.55), HAT risk (p=0.38) or bleeding risk (p=0.47) was found between the two groups. High heterogeneity was detected for thrombotic events and HAT outcomes according to I^2^.

**Figure 5.**
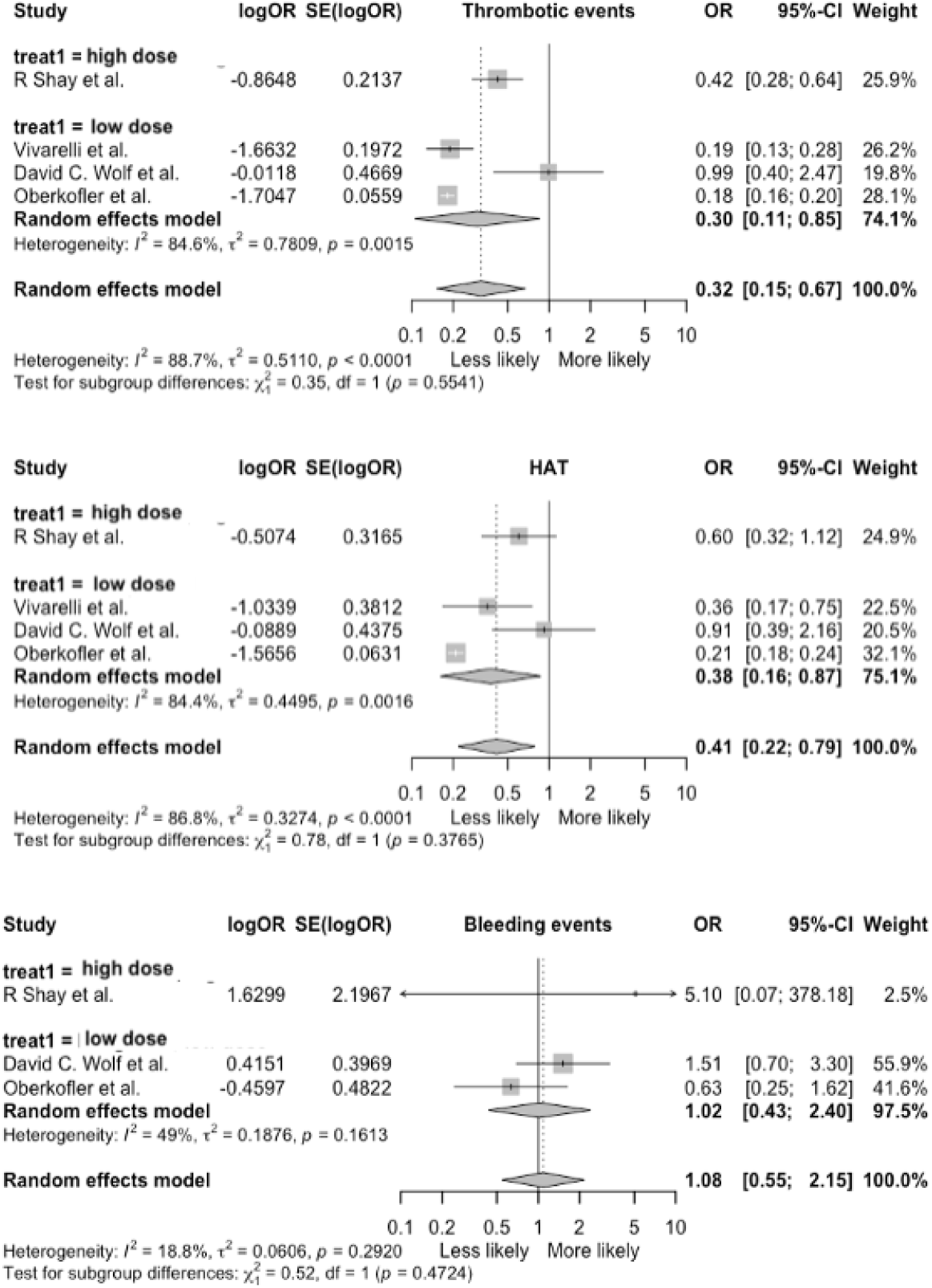
Subgroup analysis for duration of aspirin use. Studies were grouped into low-dose or high-dose. Abbreviations: OR, Hazard Ratio; CI, Confidence Interval.

Qualitative review was done for the other antithrombotic types due to either heterogeneity of dosages and durations used or the small number of studies reporting dosage or duration used that made pooling by subgroup meta-analysis impractical. The dosing and duration reported for each study is summarized in Supplemental Figure 2. For UFH, both Yip et al and Annamalai et al used 5000U subcutaneous UFH 8-hourly. For the 5 studies that reported details of UFH use that bridged to VKA therapy, warfarin was given for a range of 3 months to several years, and two studies reported an INR target of 2.0 – 3.0. For LMWH, Xie et al used a daily dose of 40mg enoxaparin until post-operative day 7. Lastly, for DOACs, Santeusanio et al used a heterogeneous mix of agents including apixaban and rivaroxaban, for a median of 351 days.

### Sensitivity analysis

A separate sensitivity analysis was performed including only randomized controlled studies and cohort studies with a high quality of evidence, which was summarized in Figure 6. The results showed that the overall ORs were largely similar to the main analysis. Moderate heterogeneity was observed in overall thrombotic events (55.1%) and HAT (31.1%); no heterogeneity was found for other outcomes.

**Figure 6.**
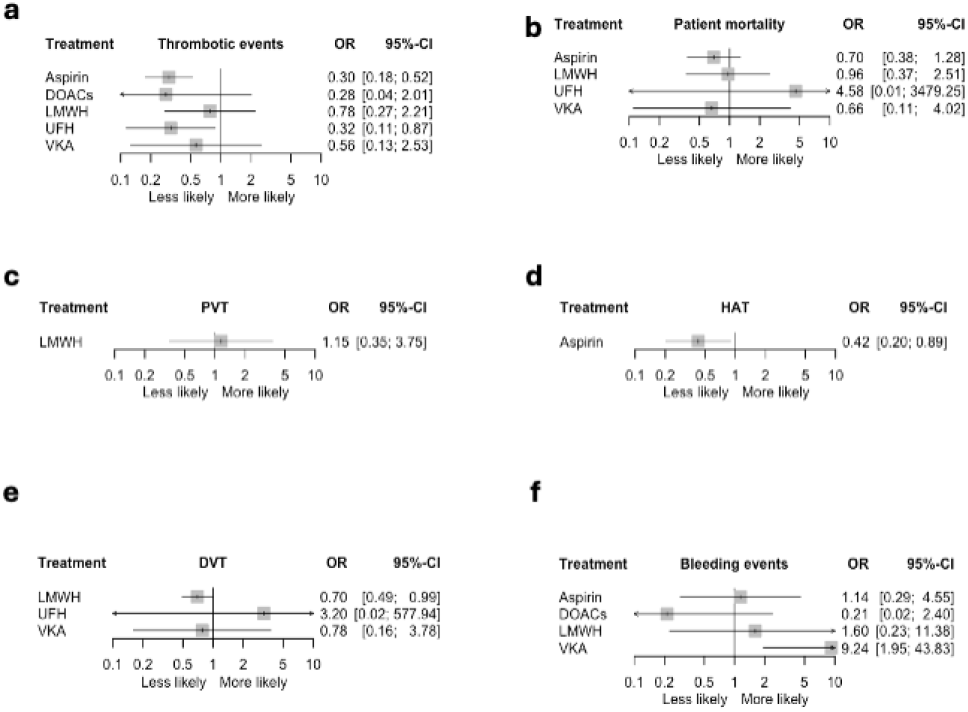
Sensitivity analysis for outcomes of study. Panels are labelled as follows: a) thrombotic events, b) patient mortality, c) portal venous thrombosis (PVT), d) hepatic artery thrombosis (HAT), e) bleeding events. No chemoprophylaxis group is the reference for all analyses. Abbreviations: OR, Hazard Ratio; CI, Confidence Interval; DOACs, Direct Oral Anticoagulants; UFH, unfractionated heparin; VKA, Vitamin K Antagonist; LMWH, Low Molecular Weight Heparin; PG, Prostaglandins and analogues.

## DISCUSSION

Thrombotic complications such as HAT, PVT, and DVT remain clinically significant concerns following liver transplantation, contributing to early graft loss, increased morbidity, and mortality. Antithrombotic strategies aim to mitigate these risks; however, their use must be carefully balanced against bleeding, especially in a postoperative liver transplant population with inherent coagulopathy and fluctuating hepatic function. Our network meta-analysis provides a comprehensive, comparative assessment of antithrombotic agents in this setting, offering new insights into the efficacy and safety of various pharmacologic options.

Our results highlight the clinically distinct management plans for arterial and venous thromboprophylaxis. With respect to arterial thromboses, aspirin demonstrated particular efficacy in preventing HAT as an antiplatelet medication. HAT remains one of the most feared early complications of LT, with incidence rates ranging from 2% to 9% and associated with poor graft outcomes^35^. Our network analysis confirms that aspirin is the best option in reducing HAT when initiated in the postoperative period, with minimal impact on bleeding risk. These findings are supported by both observational studies and clinical guidelines, which support the use of aspirin in HAT prophylaxis in the absence of contraindications^36,37^. In the subgroup analysis, no significant difference in overall thrombotic events, HAT or bleeding event risk was found between low or high dose aspirin use. As such, a more conservative approach of low-dose aspirin (75-100mg) for 3-6 months postoperatively may be warranted for HAT prophylaxis while minimising adverse effects of aspirin^37^. However, more evidence is needed to conclude on optimal aspirin dosing and duration in the post-LT context, as the small number of studies available at present limits the statistical power of this meta-analysis.

With respect to venous thromboses, heparins remain widely used in the early postoperative period^37^. Our findings demonstrate statistically significant reduction in overall thrombotic events when UFH was used, while LMWH use was linked to lower DVT risk. However, LMWH ranked second for worst associated bleeding risk and ranked lower than control for PVT risk reduction. This is in line with the results of a recent randomized controlled trial that found prophylactic enoxaparin reduced DVT risk in select subgroups of patients, but reported a notable increase in bleeding risk and no PVT risk benefit^33^. However, it is notable that this meta-analysis included studies with both high-PVT-risk populations (eg. history of PVT, thrombophilic disorders) as well as general post-LT patient cohorts. Therefore, it should be interpreted that there is little evidence for routine PVT thromboprophylaxis with heparins in every post-LT patient. However, patients with high venous thrombotic risk should consider postoperative heparin use, as LMWH was still ranked higher than all the other antithrombotic agents studied for use against PVT and DVT. This aligns with present guidelines regarding the choice of LMWH over other agents as PVT prophylaxis in high-risk PVT patients^37,38^, which may be attributable to LMWH’s more predictable anticoagulant response and reduced incidence of heparin-induced thrombocytopenia^39^.

Novel DOAC agents are a promising, yet understudied in the LT setting, alternative to traditional heparin-based therapies. In this study, DOACs ranked first for lowest bleeding risk on administration and third for reduction in overall thrombotic events. DOACs—principally apixaban and rivaroxaban—have emerged as promising options in preventing venous thromboses due to their predictable pharmacokinetics and ease of administration compared to traditional anticoagulation^8,40–42^. The results of this study are promising in highlighting DOACs as a possible new modality of post-LT thromboprophylaxis management, especially targeting venous thromboses, with lower bleeding complications and thrombotic outcomes comparable to conventional heparin-based regimens. This is consistent with retrospective cohort data in non-LT populations showing that DOACs may reduce PVT recurrence with favorable safety profiles^8,43–46^. In this study, the lack of statistical significance for DOACs can possibly be due to how few studies have been performed regarding DOAC use in the post-LT population leading to large confidence intervals after pooling. These preliminary results should encourage future prospective randomised studies on DOAC use in order to confirm these findings. In addition, as all studies on DOACs in this review included a heterogeneous mix of agents such as apixaban and rivaroxaban, studies that compare efficacy and safety of different agents, durations and dosages may inform clinical practice in the future.

Bleeding complications post-LT are particularly devastating given the technical complexity of the surgery and the altered coagulation status in these patients. In our analysis, aspirin and DOACs were associated with the lowest risk of major bleeding. This is particularly noteworthy for DOACs, as early concerns about their safety in patients with altered hepatic metabolism have led to hesitancy in their adoption^47^. However, recent pharmacokinetic studies have shown that DOACs such as apixaban are safe even in mild-to-moderate hepatic dysfunction, provided that close monitoring is ensured^47,48^. In contrast, heparin-based therapies such as LMWH, were associated with higher bleeding risks. This may reflect both their pharmacodynamic profiles and the variability in antithrombin levels and platelet function following LT^49^. Even when therapeutic levels are carefully monitored, the narrow therapeutic window of heparins poses a significant challenge in the immediate postoperative period^50^.

Vitamin□K antagonists (VKAs) are generally not recommended for thromboprophylaxis in the post–liver transplant population due to vastly increased bleeding risks postoperatively as seen from the results of this review, despite being effective at reducing in overall thrombotic risk. Furthermore, the unpredictability of hepatic metabolic function post-transplant directly affects VKA clearance and therapeutic response, making stable anticoagulant control difficult and INR targets unreliable^51^. As such, agents with predictable pharmacodynamics, shorter half-lives, and easier reversibility are more suitable in this setting.

Crucially, none of the antithrombotic agents evaluated demonstrated a statistically significant association with improved overall patient survival in our analysis. Aspirin was the best performing agent with respect to mortality, though this trend did not reach statistical significance. This is likely due to the additional effects of reducing acute cellular rejection^20^. This finding supports a targeted and tailored approach to antithrombotic use rather than routine, widespread prophylaxis, particularly given the bleeding risks associated with several agents. Prophylaxis should be individualized based on thrombotic risk factors, such as technical vascular complexity, living donor liver transplantation (LDLT), prior history of thrombosis, or the presence of hypercoagulable states. This selective strategy is in line with current consensus recommendations, which advise against universal antithrombotic use post-LT and instead recommend thromboprophylaxis in high-risk patients^37^.

### Limitations and Future Directions

Our analysis was limited by the heterogeneity of available studies and the lack of large randomized controlled trials addressing this specific question. The variation in anticoagulation protocols across studies was likely a significant source of the high heterogeneity, with differences in dosing, duration and time of initiation varying from study to study. We attempted to address this via the subgroup analysis of dosages and durations. Another likely source of heterogeneity was the patient cohorts, with some studies studying all patients with normal thrombotic risk while others only anticoagulating patients with history of thrombosis or other risk factors, or even according to individual physician discretion. This is likely a reason why results varied between studies as patients had vastly different risk profiles, but we were unable to meaningfully subgroup them due to either small number of studies, the regimens used being too heterogeneous or reporting being unclear for some.

Secondly, most studies were not randomized in design, compromising the power of our results as the baseline characteristics of the thromboprophylaxis versus control group in some studies were not strictly comparable. The inclusion of cohort studies and small cohort sizes may also bias the results. However, major evidence and randomized controlled trials on this topic remains relatively scarce, and performing a comprehensive analysis and comparison of the numerous thromboprophylaxis methods in the post-LT setting relying only on RCTs is difficult at the present. Specifically, further investigation is needed regarding the role and safety of newer antithrombotic agents, especially DOACs.

Future research should focus on prospective, randomized studies that can more definitively establish the optimal thromboprophylaxis strategies for different patient populations, possibly with a focus on DOACs, aspirin and LMWH in line with the conclusions of this meta-analysis as well as current literature. Specifically, studies comparing specific DOAC agents such as apixaban and rivaroxaban in the post-LT setting could be informative given that DOACs appear promising for post-LT thromboprophylaxis.

## Conclusion

In conclusion, our analysis quantitatively highlights aspirin and DOACs as the most promising thromboprophylaxis options following liver transplantation, offering a meaningful reduction in thrombotic complications with the lowest associated bleeding risk, while also providing a comparison and recommendation between dosing regimens and duration of use. Aspirin is specifically suitable for arterial thromboprophylaxis including HAT, while DOACs appear to be a promising option in preventing venous thromboses such as PVT and DVT over conventional LMWH therapy, especially in the high-risk population. However, no antithrombotic agent demonstrated a statistically significant survival benefit in the general LT population, likely due to balancing with an increased bleeding risk, suggesting that routine prophylaxis for all post-LT recipients is not warranted. Instead, antithrombotic strategies should be reserved for selected high-risk populations, guided by clinical and technical risk factors, and aligned with evolving international recommendations. Future prospective, controlled studies are needed to further define the optimal agent, duration, and patient selection criteria.

## Data Availability

All data is acquired from public databases Medline and Embase.

## List of Abbreviations

LT: Liver Transplantation
HAT: Hepatic Artery Thrombosis
PVT: Portal Vein Thrombosis
DVT: Deep Vein Thrombosis
UFH: Unfractionated Heparin
LMWH: Low Molecular Weight Heparin
VKA: Vitamin K Antagonists
DOACs: Direct Oral Anticoagulants
OR: Odds Ratio
CI: Confidence Interval

## DECLARATIONS

## Acknowledgements

No additional acknowledgements.

## Sources of Funding

No funding was required for this study.

## Authors’ Contributions

All authors approve the final version of the manuscript, including the authorship list and agree to be accountable for all aspects of the work in ensuring that questions related to the accuracy or integrity of any part of the work are appropriately investigated and resolved. All authors have made substantial contributions to the following: (1) the conception and design of the study, or acquisition of data, or analysis and interpretation of data, (2) drafting the article or revising it critically for important intellectual content, (3) final approval of the version to be submitted. No writing assistance was obtained in the preparation of the manuscript.

The manuscript, including related data, figures and tables has not been previously published and that the manuscript is not under consideration elsewhere.

**Conceptualisation and Design:** AK, JHL

**Acquisition of Data:** CEYO, RSYW, YZ, YDC, YZN

**Analysis and Interpretation of Data:** CEYO, AK, JHL

**Writing – original draft:** CEYO, RSYW, YZ, YDC, YZN

**Writing – review & editing:** AK, JHL, CEYO

## Conflicts of Interest

The authors declare that they have no competing interests.

## Ethical Statement

Not applicable. The study was conducted in accordance with the Declaration of Helsinki. The study was exempt from IRB review was no confidential patient information was involved.

## Transparency Statement

AK affirms that the manuscript is an honest, accurate, and transparent account of the study being reported; that no aspects of the study have been omitted; and that any discrepancies from the study as originally planned (and, if relevant, registered) have been explained.

## TABLES AND FIGURES

(see Supplemental Figures.pdf)

**Supplemental Figure 1**. Search strategy for Medline and Embase

**Supplemental Figure 2**. Summary of included studies

**Supplemental Figure 3**. Risk of bias in randomized controlled trials

**Supplemental Figure 4**. GRADE Working Group grades of evidence

**Supplemental Figure 5**. Test for consistency

## Notes

### Competing Interest Statement

The authors have declared no competing interest.

### Funding Statement

No funding was obtained.

