## Supplemental Figures for "Comparison of Efficacy, Safety, and Survival Outcomes of Anticoagulation and Antiplatelet Strategies Post Liver Transplantation: A Systematic Review and Network Meta-Analysis"

**Supplemental Figure 1: Search strategy for Medline and Embase**

Medline

1. Liver Transplantation/ or Organ Transplantation/ or (liver transplant$).tw or (organ transplant$).tw
2. exp Anticoagulants/ or anticoag$.tw or (heparin or UFH or LMWH or dalteparin or enoxaparin or Clexane or fondaparinux).tw or ((oral anticoagula$) or DOAC or NOAC or TSOAC or dabigatran or rivaroxaban or apixaban or edoxaban).tw or (warfarin or coumarin or (vitamin K antagonist$)).tw or ((AT III) or ATIII or antithrombin).tw or ((direct thrombin inhibitor$) or argatroban or desirudin or bivalirudin).tw
3. Platelet Aggregation Inhibitors/ or aspirin.tw or ((adenosine diphosphate receptor inhibitor$) or (ADP receptor inhibitor$) or clopidogrel or ticlopidine or ticagrelor or prasugrel).tw or ((adenosine reuptake inhibitor$) or dipyridamole).tw or ((glycoprotein platelet inhibitor$) or abciximab or eptifibatide or tirofiban).tw or ((phosphodiesterase inhibitor$) or cilostazol).tw or ((PAR-1 antagonist$) or (protease-activated receptor antagonist$) or vorapaxar).tw or antiplatelet$.tw or (prostaglandin$ or iloprost or PG$).tw
4. 2 or 3
5. 1 and 4

Embase

1. 'liver transplantation'/exp OR 'liver transplantation' OR 'liver graft':ti,ab OR 'liver transplant*':ti,ab
2. 'anticoagulant agent'/exp OR 'anticoagulation'/exp/mj OR 'anticoag*':ti,ab OR 'heparin':ti,ab OR 'UFH':ti,ab OR 'LWMH':ti,ab OR 'dalteparin':ti,ab OR 'enoxaparin':ti,ab OR 'Clexane':ti,ab OR 'fondaparinux':ti,ab OR 'oral anticoag$':ti,ab OR 'DOAC':ti,ab OR 'NOAC':ti,ab OR 'TSOAC':ti,ab OR 'dabigatran':ti,ab OR 'rivaroxaban':ti,ab OR 'apixaban':ti,ab OR 'edoxaban':ti,ab OR 'warfarin':ti,ab OR 'coumarin':ti,ab OR 'vitamin K antagonist$':ti,ab OR 'AT III':ti,ab OR 'ATIII':ti,ab OR 'antithrombin':ti,ab OR 'direct thrombin inhibitor$':ti,ab OR 'argatroban':ti,ab OR 'desirudin':ti,ab OR 'bivalirudin':ti,ab
3. 'antiplatelet activity'/exp OR 'aspirin':ti,ab OR 'adenosine diphosphate receptor inhibitor$':ti,ab OR 'ADP receptor inhibitor$':ti,ab OR 'clopidogrel':ti,ab OR 'ticlopidine':ti,ab OR 'ticagrelor':ti,ab OR 'prasugrel':ti,ab OR 'adenosine reuptake inhibitor$':ti,ab OR 'dipyridamole':ti,ab OR 'glycoprotein platelet inhibitor$':ti,ab OR 'abciximab':ti,ab OR 'eptifibatide':ti,ab OR 'tirofiban':ti,ab OR 'phosphodiesterase inhibitor$':ti,ab OR 'cilostazol':ti,ab OR 'PAR-1 antagonist$':ti,ab OR 'protease-activated receptor antagonist$':ti,ab OR 'vorapaxar':ti,ab OR 'antiplatelet$':ti,ab OR 'prostaglandin$':ti,ab OR 'iloprost':ti,ab OR 'PG$':ti,ab
4. #2 OR #3
5. #1 and #4
6. #1 AND #4 AND [english]/lim AND [humans]/lim

**Supplemental Figure 2: Summary of included studies**

| **Author** | **Year** | **Country** | **Study design** | **Sample size** | **Intervention(s)** | **Mean age (years)** | **Quality assessment** |
| --- | --- | --- | --- | --- | --- | --- | --- |
| Shay et al. | 2013 | United States | Retrospective cohort | 469 | Aspirin 325mg orally daily for 3 months, from POD0 | 52.06 | 8 |
| Vivarelli et al. | 2007 | United States | Retrospective cohort | 828 | Aspirin 100mg per day from POD8 | NR | 11 |
| Wolf et al. | 1997 | United States | Retrospective cohort | 529 | Aspirin 81mg daily from POD1 | NR | 8 |
| Oberkofler et al. | 2022 | International multicentre study | Retrospective cohort | 2351 | Aspirin 75-100mg daily indefinitely | 54.88 | 9 |
| Chung et al. | 2024 | Australia | Retrospective cohort | 311 | Aspirin | 58.68 | 8 |
| Yip et al. | 2016 | United States | Retrospective cohort | 999 | UFH subcutaneously 5000U 8-hourly | NR | 8 |
| Annamalai et al. | 2014 | United States | Retrospective cohort | 314 | UFH 5000 U | 55.08 | 10 |
| Salami et al. | 2013 | United States | Retrospective cohort | 920 | UFH and warfarin for 3-6months | 49.95 | 8 |
| Bos et al. | 2021 | Netherlands, France | Retrospective cohort | 235 | UFH and warfarin, INR target 2.0-3.0, 3 months duration | 51.75 | 7 |
| Widén et al. | 2009 | United Kingdom | Retrospective cohort | 138 | UFH and warfarin, mean duration 3.15 years | 47.96 | 9 |
| Hu et al. | 2022 | China | Retrospective cohort | 33 | UFH and warfarin, median duration 17.6 months | 54.74 | 5 |
| Santeusanio et al. | 2020 | United States | Retrospective cohort | 40 | DOACs compared against UFH and warfarin | NR | 8 |
| Xie et al | 2025 | China | RCT | 462 | LMWH 40mg daily, POD1-7 | 52.00 | **-*** |

*** see Supplemental Figure for evaluation of RCTs. Legend:** LMWH, Low Molecular Weight Heparin; UFH, Unfractionated Heparin; DOACs, Direct Oral Anticoagulants; POD, Post-operative Day; RCT, Randomised Controlled Trial; NR, not reported.

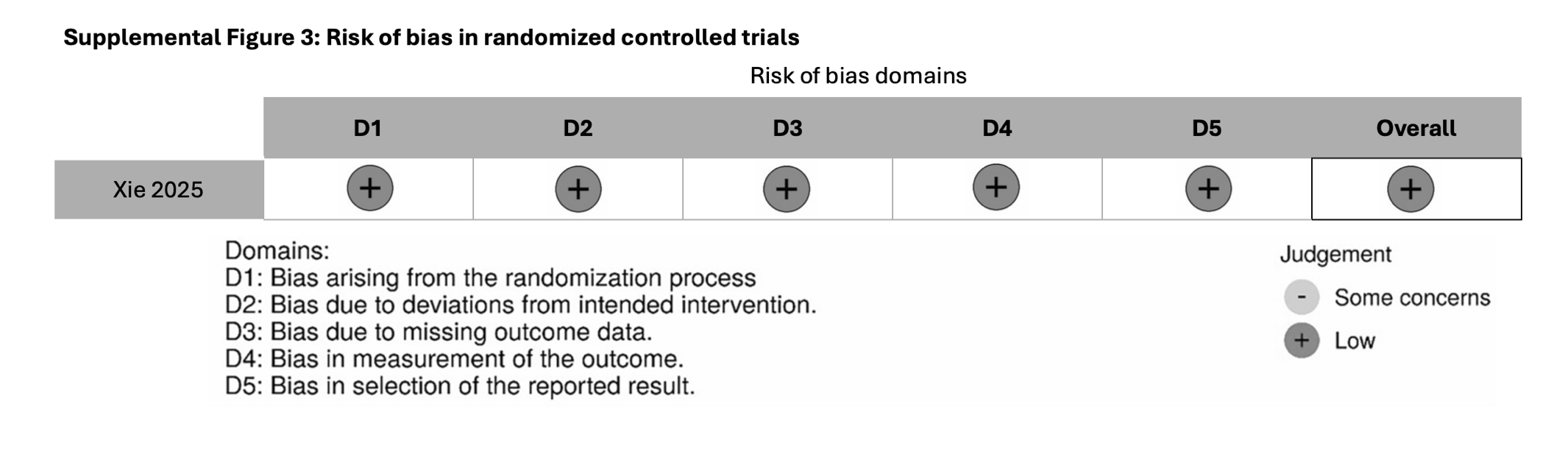

**Supplemental Figure 4: GRADE Working Group grades of evidence**

| **Outcome** | **Certainty of Evidence** | **Comments** |
| --- | --- | --- |
| **Thrombotic events** |  |  |
| Control vs Aspirin | ⊕⊕OO  **Low** | Low certainty of evidence due to indirectness (first-order loop via direct comparisons with No Treatment) |
| Control vs DOACs | ⊕⊕OO  **Low** | Low certainty of evidence due to risk of bias, indirectness (first-order loop via direct comparisons with No Treatment) |
| Control vs UFH | ⊕⊕OO  **Low** | Low certainty of evidence due to risk of bias, indirectness (first-order loop via direct comparisons with No Treatment) |
| Control vs VKA | ⊕⊕OO  **Low** | Low certainty of evidence due to indirectness (first-order loop via direct comparisons with No Treatment) |
| Control vs LMWH | ⊕⊕OO  **Low** | Low certainty of evidence due to risk of bias |
| **Patient mortality** |  |  |
| Control vs Aspirin | ⊕⊕OO  **Low** | Low certainty of evidence due to indirectness (first-order loop via direct comparisons with No Treatment) and risk of bias. |
| Control vs UFH | ⊕⊕OO  **Low** | Low certainty of evidence due to indirectness (first-order loop via direct comparisons with No Treatment) |
| Control vs VKA | ⊕OOO  **Very Low** | Very low certainty of evidence due to risk of bias and imprecision (large confidence intervals) |
| Control vs LMWH | ⊕⊕OO  **Low** | Low certainty of evidence due to indirectness (first-order loop via direct comparisons with No Treatment) and risk of bias. |
| **PVT** |  |  |
| Control vs VKA | ⊕OOO  **Very Low** | Very low certainty of evidence due to risk of bias and imprecision (large confidence intervals) |
| Control vs LMWH | ⊕⊕OO  **Low** | Low certainty of evidence due to risk of bias |
| **HAT** |  |  |
| Control vs Aspirin | ⊕⊕OO  **Low** | Low certainty of evidence due risk of bias and imprecision (large confidence intervals) |
| Control vs VKA | ⊕⊕OO  **Low** | Low certainty of evidence due to risk of bias, indirectness (first-order loop via direct comparisons with No Treatment) |
| **DVT** |  |  |
| Control vs UFH | ⊕⊕OO  **Low** | Low certainty of evidence due to risk of bias, indirectness (first-order loop via direct comparisons with No Treatment) |
| Control vs VKA | ⊕⊕OO  **Low** | Low certainty of evidence due to risk of bias, indirectness (first-order loop via direct comparisons with No Treatment) |
| Control vs LMWH | ⊕⊕OO  **Low** | Low certainty of evidence due to risk of bias |
| **Bleeding events** |  |  |
| Control vs Aspirin | ⊕⊕OO  **Low** | Low certainty of evidence due to risk of bias, indirectness (first-order loop via direct comparisons with No Treatment) |
| Control vs DOACs | ⊕⊕OO  **Low** | Low certainty of evidence due to risk of bias, indirectness (first-order loop via direct comparisons with No Treatment) |
| Control vs UFH+VKA | ⊕⊕OO  **Low** | Low certainty of evidence due to risk of bias, indirectness (first-order loop via direct comparisons with No Treatment) |
| Control vs LMWH | ⊕OOO  **Very Low** | Very low certainty of evidence due to risk of bias and imprecision (large confidence intervals) |

**Abbreviations:** PIM, Portal Inflow Modulation; SAL, Splenic Artery Ligation; PCS, Portocaval Shunt.

**GRADE Working Group grades of evidence**

**High quality:** Further research is very unlikely to change our confidence in the estimate of effect.

**Moderate quality:** Further research is likely to have an important impact on our confidence in the estimate of effect and may change the estimate.

**Low quality:** Further research is very likely to have an important impact on our confidence in the estimate of effect and is likely to change the estimate.

**Very low quality:** We are very uncertain about the estimate.

**Supplemental Figure 5: Test for consistency**

|  | | | **Q** | **p-value** |
| --- | --- | --- | --- | --- |
| Thrombotic events | | |  |  |
|  | Control vs LMWH | | 5.98 | 0.05 |
|  | Control vs UFH+VKA | | 5.81 | 0.05 |
|  | Control vs aspirin | | 5.02 | 0.08 |
|  | DOACs vs UFH+VKA | | 0.02 | 0.88 |
| Patient mortality | | |  |  |
|  | | Control vs LMWH | 0.20 | 0.66 |
| DVT | | |  |  |
|  | | Control vs LMWH | 0.08 | 0.77 |
| PVT | | | | |
|  | | Control vs LMWH | 0.95 | 0.62 |
| HAT | | | | |
|  | | Control vs aspirin | 4.35 | 0.23 |
| Bleeding events | | | | |
|  | | Control vs LMWH | 0.48 | 0.49 |
|  | | Control vs UFH+VKA | 0.85 | 0.58 |
|  | | Control vs aspirin | 0.46 | 0.79 |
|  | | DOACs vs UFH+VKA | 0.03 | 0.87 |

Legend: * bolded p-value ≤0.05 denotes statistical significance. Abbreviations: LMWH, Low Molecular Weight Heparin; UFH, Unfractionated Heparin; VKA, Vitamin K Antagonist; DOACs, Direct Oral Anticoagulant.
